# Dissecting the functional landscape of rare diseases through genomic variation in a heterogeneous cohort of 11,000 patients

**DOI:** 10.64898/2026.06.10.26355349

**Authors:** Graciela Uria-Regojo, Lidia Fernández-Caballero, Aroa López-Alcojor, Lucia Lopez-Lopez, Yolanda Benitez, Cristina Rodilla, Almudena Avila-Fernandez, María José Trujillo-Tiebas, Ana Osorio, Marta Corton, Berta Almoguera, Carmen Ayuso, Pablo Minguez

## Abstract

Rare diseases (RDs) pose a major diagnostic challenge. Genetic and phenotypic heterogeneity, incomplete knowledge of disease mechanisms, and limitations in variant clinical interpretation leave many patients without a molecular diagnosis. Meanwhile, the growing volume of genomic data generated in clinical practice offers an opportunity to develop data-driven methodologies for exploring disease mechanisms and supporting the reanalysis of unsolved cases. In this study, we aggregated real-world genomic data from 11,084 unrelated patients clinically classified into 122 diseases. We built a multi-disease genomic variant frequency database (FJD-DB), which enabled the development of variant and gene-disease association scores by means of case-control subcohort comparisons across 32 disease groups. Functional enrichment analyses were then used to highlight disease-associated protein domains, pathways, biological processes, and phenotypes. Finally, the resulting knowledge was integrated into a data-driven framework for the guided reanalysis of unsolved RD patients applied to Inherited Retinal Dystrophies (IRD) patients as first use case.

The FJD-DB contained over 45 million unique variants, including ∼184,000 potentially pathogenic variants. We identified disease-associated pathogenic variants and highlighted both established and candidate disease genes. We identified 179 protein domains, 124 Human Phenotype Ontology terms, 79 Reactome pathways, and 72 Gene Ontology biological processes significantly enriched across multiple diseases, revealing disease-specific functional signatures. Integration of disease-associated variant, gene, and functional signals enabled the development of a data-driven framework for guided reanalysis of unsolved RD cases. Applied to over 1,100 unsolved IRD cases, this data-driven reanalysis yielded clinically relevant findings in 32 patients, including three molecular diagnoses, 25 possibly solved cases, and four cases in which prioritized variants highlighted four candidate genes. We illustrate how aggregated real-world genomic data can be leveraged to identify disease-associated molecular signals generating novel biological hypotheses. A unified analytical framework provides a scalable strategy for knowledge discovery and guided reanalysis, facilitating the identification of overlooked and potentially novel genetic causes of RDs.

## BACKGROUND

Rare diseases (RDs) affect more than 300 million people worldwide and are predominantly genetic in origin [1]. Despite the widespread implementation of high-throughput sequencing techniques, a substantial proportion of patients remain without a molecular diagnosis due to extensive genetic and phenotypic heterogeneity, the complexity of human variation architecture, and limitations in current variant interpretation frameworks [2–4]. Consequently, many patients undergo prolonged diagnostic journeys that can extend for years before reaching a diagnosis [5].

Periodic and systematic reanalysis has emerged as an effective strategy to improve diagnostic yield in unsolved cases [6]. The continuous discovery of novel disease genes, the reclassification of genetic variants, and the development of improved bioinformatic approaches have enabled the identification of diagnoses that were missed during the initial assessments [7–9]. At the same time, the increasing adoption of genomic sequencing in clinical practice has generated large collections of real-world data that remain largely underexploited beyond their primary diagnostic purpose. These datasets encompass heterogeneous patients, phenotypes, sequencing strategies, and diagnostic outcomes, capturing the true complexity of routine clinical genetics [10–12]. While this heterogeneity introduces analytical challenges, it also creates an unprecedent opportunity to maximize the value of genomic data already generated during the diagnostic process [13,14].

Genetic variants causing similar phenotypes frequently disrupt common biological functions, either by affecting the same genes or by altering genes involved in related pathways, molecular processes, protein complexes, or cellular structures [15–17]. This functional convergence has been widely exploited in gene discovery approaches based on guilt-by-association principles, where novel disease genes are prioritized according to their functional similarity to previously established disease-causing genes [18–20].

Motivated by the need for new approaches to boost genetic diagnostic rates in RDs, we aggregated real-world genomic data generated through both diagnostic and research activities from a large heterogeneous cohort of over 11,000 RD patients and analyzed them using a unified bioinformatic framework. By combining variant, gene, and functional association analyses, we systematically explored the functional landscape of more than 120 RDs, uncovering association signals at the level of variants, genes, protein domains, biological processes, pathways, and phenotypes. This approach enabled the generation of novel genotype-phenotype hypotheses and the proposal of candidate disease mechanisms across disorders. Finally, we integrated the resulting knowledge into a data-driven diagnostic framework and prioritization algorithms designed to support the guided reanalysis of unsolved cases, particularly those in which the underlying genetic cause has remain unknown.

## METHODS

### Cohort Description and Development of a Curated Disease Ontology

We retrospectively selected all cases with next generation sequencing (NGS) tests performed at the Genetics and Genomics Department of the University Hospital Fundación Jiménez Díaz (UH-FJD, Madrid, Spain) from September 2015 to June 2024. Overall, 11,084 unrelated patients were included in the cohort of study. Samples were sequenced using a range of sequencing tests: 1) Commercial sequencing panels (N = 10,604): TruSightOne Sequencing Panel kit (TSO, Illumina, San Diego, CA, USA) and Clinical Exome Solution Sequencing Panel kit versions 1, 2 and 3 (CES, Sophia Genetics, Boston, MA, USA), targeting 4,813, 4,828 and 5,287 genes, respectively; 2) Whole Exome Sequencing (WES) using Agilent SureSelect v6 (Agilent Technologies, Santa Clara, CA, USA) or the Twist Human Core Exome (Twist Bioscience) libraries (N = 354); and 3) Whole Genome Sequencing (WGS) (N = 126).

Patients were clinically classified according to their suspected disease by the molecular geneticists of the UH-FJD Genetics Department using a hierarchical three-level classification framework. The top-level (Level-1) comprised 19 general disease categories (e.g. Ophthalmic diseases, Cardiopathies and circulatory system disorders, Oncohematology), which are further subdivided in 41 groups of related diseases (Level-2) (e.g. Inherited Retinal Dystrophies, Corneal Dystrophy, Ocular Malformations), and finally into 122 specific diseases at Level-3 (e.g. Albinism, Cataracts, Anterior Segment Dysgenesis), as specified in Supplementary Table S1. All patients were classified at the three hierarchical levels and could potentially be assigned to multiple cohorts at the same hierarchical level.

To perform comparative analysis, we defined a control cohort (pseudocontrol) for each subcohort derived from the different classes. Each pseudocontrol cohort includes all patients from the FJD-DB who do not suffer from any disease falling within the top-level (Level 1) disease category containing the specific disease of interest.

### Variant detection analysis

All sequencing data were analyzed following the GATK best practices [21]. Data from 6,688 patients (60.3% of the overall cohort) were analyzed as previously described by Iancu et al.[10] and converted from GRCh37/hg19 to GRCh38/hg38 using a liftover process. The remaining samples (N = 4396; 39.7%) were processed using the GRCh38/hg38 genome assembly with an updated version of the same pipeline (https://github.com/TBLabFJD/PARROT-FJD). The entire cohort was annotated using PARROT-FJD (https://github.com/TBLabFJD/PARROT-FJD/blob/main/PARROT-FJD_annotation.xlsx).

### Construction of the Multi-Disease Local Database of Genomic Variant Frequencies

We constructed a local database of allele frequencies (FJD-DB) as described by Iancu et al. [10]. For patients or samples with multiple sequencing tests, we kept the test with the highest target coverage prioritizing WGS over WES, WES over CES, and a CES with a larger padded target region over one with less covered positions. We discarded patient relatives by using PLINK [22]. After sample filtering, we merged the remaining samples (N = 11,084) into a multi-sample vcf file. Then, to differentiate between true non-mutated positions and those not covered we used sequencing coverage information to impute variant genotypes as reference homozygotes (0/0) at sites covered by more than 10 reads in each patient. We calculated allele frequencies for the whole cohort, and for each of the 182 individual subcohorts: 19 Level-1 categories, 41 Level-2 diseases and 122 Level-3 specific diseases, as well as for their corresponding pseudocontrol cohorts, using *in-house* scripts based on the Hail python library (https://hail.is). The code for the database construction is available at https://github.com/TBLabFJD/DBofAFs.

We restricted subsequent analyses to 32 subcohorts defined by Level-2 disease categories after excluding categories with small sample sizes (N < 15), as well as healthy or clinically ambiguous groups (e.g. Congenital Anomalies and Various Cancer) (Supplementary Table S1).

### Variant Disease Association Score

To estimate the level of association between a genetic variant and a given disease category, we developed the Variant Disease Association Score (VDAS). This score is a cohort-specific metric and considers two ratios comparing each disease-category cohort with its corresponding pseudocontrol cohort: 1) the allele frequency ratio and 2) the ratio of the number of homozygous cases. The VDAS is defined as follows:

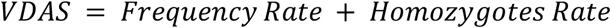

from which

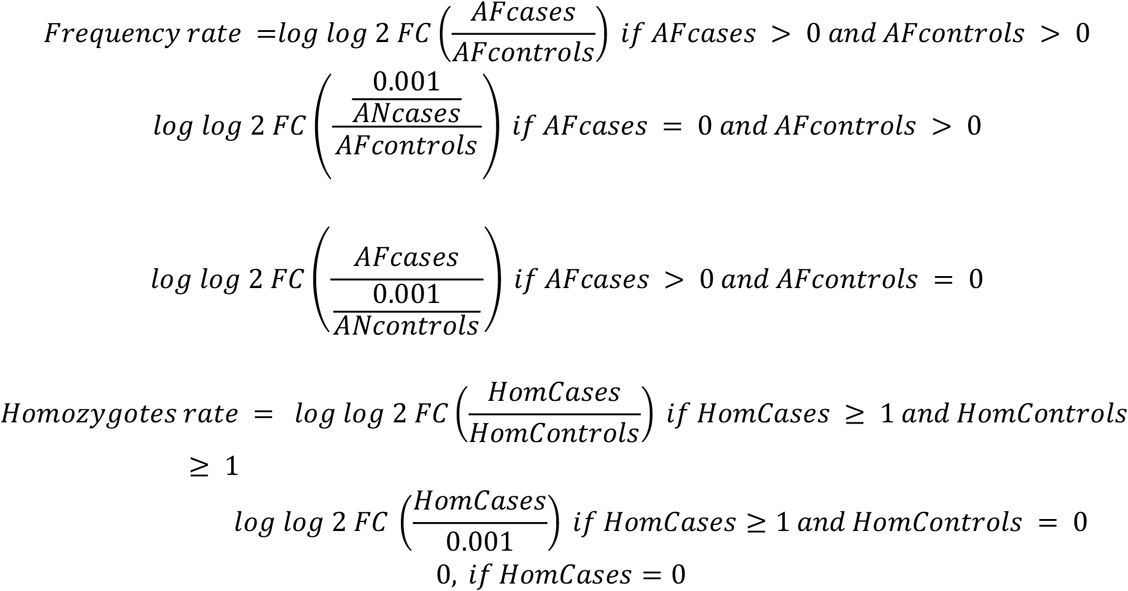

Being:

- *AFcases*, the variant’s allele frequency in the disease cohort.
- *AFcontrols*, the variant’s allele frequency in the pseudocontrol cohort.
- *ANcases*, the variant’s allele number in the disease cohort.
- *ANcontrols*, the variant’s allele frequency in the pseudocontrol cohort.
- *HomCases*, the number of homozygote patients in the disease cohort carrying the variant.
- *HomControls*, the number of homozygote pseudocontrols in the pseudocontrol cohort carrying the variant.

Thus, the VDAS was applied to subcohorts defined by our Level-2 disease classification. For each Level-2 group, the VDAS was calculated for all pathogenic or predicted pathogenic genetic variants (hereafter collectively referred to as pathogenic variants) present in the FJD-DB and covered in both the disease and pseudocontrol cohorts (*ANcases* > 0 and *Ancontrols* > 0). We define pathogenic variants as those meeting at least one of the following criteria: i) annotated in ClinVar [23] as “Pathogenic” or “Likely Pathogenic”; ii) CADD_PHRED ≥ 25 [24]; iii) Variant consequence classified as frameshift or stop-gained. Conversely, variants annotated in ClinVar as “Benign” or “Likely Benign” were excluded, regardless of any other criteria met.

The homozygotes rate was calculated only for variants located on autosomal chromosomes (chr. 1-22); for variants on sex chromosomes (X and Y), this value was set to 0.

### Enrichment analysis of protein domains across disease-specific VDAS distributions

We aimed to identify protein domains enriched in disease-associated pathogenic variants using gene set enrichment analysis (GSEA; [25]). For each disease, VDAS values were transformed into normalized VDAS (nVDAS) as follows:

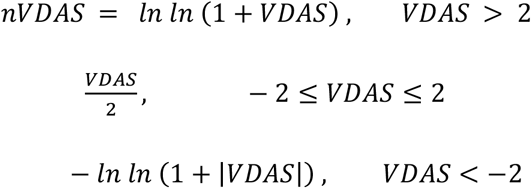

This transformation preserves the directionality of the score while compressing extreme values and reducing the large gaps between the main VDAS groups.

For each Level-2 disease category, we applied GSEA to variants ranked by nVDAS values. As the annotation set, we used protein domains from the InterPro database [26], which were assigned to variants based on their position within MANE transcripts. To prioritize protein domains truly enriched in pathogenic variants associated with cases, and to minimize potential artifacts derived from the ranking distribution, we considered protein domains statistically significant when they met all the following criteria: i) false discovery rate (FDR) ≤ 0.25, ii) normalized enrichment score (NES) > 0, and iii) at least 75% of the core enrichment pathogenic variants had a nVDAS > 0.

### Gene Disease Association Score

To estimate the level of association between a gene and a genetic disease, we developed the Gene Disease Association Score (GDAS). Similar to the VDAS, the GDAS is a cohort-specific metric calculated independently for each gene and disease cohort as follows:

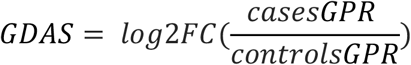

where:

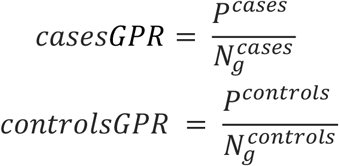

Being:

- *casesGPR*, the Gene Pathogenicity Ratio in the disease cohort.
- *P*^*cases*^, the number of occurrences of pathogenic variants within the gene coordinates, considering both heterozygous and homozygous states. This included: i) variants observed exclusively in patients, and ii) variants among the top 10% highest VDAS-scoring variants detected in both patients and controls, including all variants tied at the cutoff score. When Pcases = 0, it was set to 0.001.
- 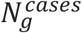,the number of patients (cases) with sequencing data covering a specific gene *g*. Given differences in coverage across CES, WES, and WGS, this value is defined per gene as follows: genes included in the CES panel used the full cohort of patients; protein-coding genes not included in CES used patients with WES or WGS data; and non-protein-coding genes used only patients with WGS data.
- *controlsGPR*, the Gene Pathogenicity Ratio in the pseudocontrol cohort.
- *P*^*controls*^, the number of occurrences of pathogenic variants within the gene coordinates observed exclusively in pseudocontrols, considering both heterozygous and homozygous states. When Pcontrols = 0, it was set to 0.001.
- 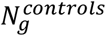, the number of pseudocontrols with sequencing data covering a specific gene *g*. Given differences in coverage across CES, WES, and WGS, this value is defined per gene as follows: genes included in the CES panel used the full cohort of pseudocontrols; protein-coding genes not included in CES used pseudocontrols with WES or WGS data; and non–protein-coding genes used only pseudocontrols with WGS data.

Thus, the GDAS was applied to subcohorts defined by our Level-2 disease classification.

### Identification of genes with the highest pathogenic mutational burden across diseases

We aimed to identify the genes showing a high pathogenic mutational burden for each disease within the Level-2 classification. To this end, we first removed genes with pathogenic variants showing very low occurrence counts in each disease:

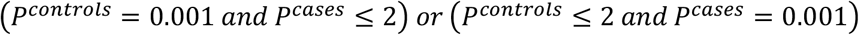

Furthermore, for diseases with no patients sequenced by WES or WGS, only genes covered in the CES were kept, for the rest of the diseases only protein-coding genes were kept.Then, we selected genes with a GDAS ≥ 3 and ranked them according to their *Pcases* value to prioritize representative genes with stronger disease-association signals for visualization and discussion purposes. These selected genes were termed High Pathogenic Burden Genes (HPBGs) and were further classified into two categories: i) genes previously associated with the disease according to the ORPHA database[27], or ii) genes not currently associated with the disease in the ORPHA database. Correspondences between ORPHA terms and Level-2 diseases were manually curated (Supplementary Table S2). For inherited retinal dystrophies (IRDs), the *RS1* gene was included as a gene previously associated with the disease [28], even though it lacked a corresponding ORPHA term.

### Enrichment analysis of biological processes, pathways, and human phenotypes across disease-specific GDAS distributions

We aimed to identify biological processes, molecular pathways, and human phenotypic features showing a significant enrichment of genes carrying a high burden of pathogenic variants associated with each disease. Like the VDAS, the GDAS values were normalized prior to GSEA as follow:

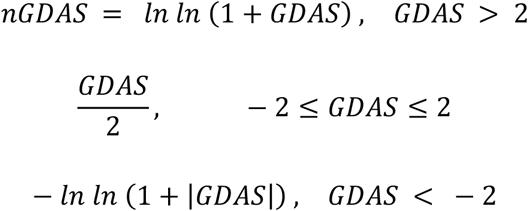

Following nGDAS normalization, we selected the subset of genes to be used as input for a GSEA analysis for each Level-2 disease. Genes with pathogenic variants showing very low occurrence counts:

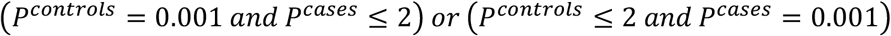

were removed from the enrichment analysis to reduce instability driven by sparsely represented genes. For diseases with no patients sequenced by WES or WGS, only genes covered in the CES were analyzed, for the rest of the diseases only protein-coding genes were analyzed. As annotation set, we used: i) Biological process terms from the Gene Ontology (GO) database [29], ii) Biological pathway terms from Reactome [30] and KEGG [31], and iii) Human phenotype terms from the Human Phenotype Ontology (HPO) database [32]. We extracted biological processes, pathways and human phenotypes that met all the following criteria: i) normalized enrichment score (NES) > 0, ii) false discovery rate (FDR) ≤ 0.25, and iii) at least 75% of the core enrichment genes had a nGDAS > 0. As with protein domains, this additional filtering step was included to ensure that the identified enrichments truly reflect genes with a high pathogenic mutational burden associated with cases, rather than artifacts derived from the ranked distribution.

### A Prioritization Algorithm for the Reanalysis of Unsolved Inherited Retinal Dystrophies cases

We developed a disease-specific, data-driven prioritization algorithm integrating variant-level associations (VDAS), gene-level pathogenic burden (GDAS), and enriched biological functions to prioritize candidate variants in unsolved cases. As a first use case, the algorithm was applied to IRDs. Starting from all variants from the FJD-DB observed in patients with the selected phenotype or disease (IRD in our first use case), we applied a stepwise filtering strategy to prioritize potentially relevant candidates.

First, we retained variants classified as “Pathogenic” or “Likely Pathogenic” in ClinVar. For variants of uncertain significance (VUS), conflicting interpretation, or lacking a ClinVar annotation, only rare (gnomAD AF_popmax ≤ 0.01; [13]) were kept. In addition, we selected those fulfilling at least one of the following criteria: i) CADD_PHRED ≥ 15, ii) frameshift or stop-gained consequence, iii) SpliceAI_max ≥ 0.25 [33] affecting Intronic positions or iv) SpliceAI_max ≥ 0.25 affecting Exonic Synonymous positions.

Next, the VDAS was calculated for all selected variants, retaining only those with a positive disease association signal (VDAS > 0), defined as having a higher burden in cases compared to pseudocontrols, and present in unsolved cases of the IRD subcohort. These remaining variants were then prioritized in three levels of gene association with IRD:

a. in genes with prior evidence, including genes present in at least one of the following resources: a) the PanelApp Retinal Disorders Green or Amber lists (https://nhsgms-panelapp.genomicsengland.co.uk/panels/307/v7.0), b) Red (candidate) genes in the same Retinal Disorders PanelApp panel; c) genes proposed by the European Retinal Disease Consortium (ERDC; https://www.erdc.info/candidate-ird-genes); and d) internal candidate genes currently under follow-up in our laboratory.
b. in data-supported candidate genes; defined as genes fulfilling all the following criteria: a) GDAS-IRD > 0; b) retinal expression detected in the GTEx database [34] or having unavailable expression data (RNA expression = NA), and c) gene is part of the core-enrichment set of genes annotated to at least one IRD-enriched functional term (Gene Ontology, Reactome. KEGG or HPO) identified in the previous enrichment analyses.
c. in genes without prior evidence or other genome regions, including all remaining genes not assigned to the previous groups. To prioritize these genes, we integrated their GDAS with the predictive score provided by GLOWgenes [35], a gene-disease association predictor applied to IRD. The complete GLOWgenes IRD ranking is available at https://github.com/TBLabFJD/GLOWgenes/blob/master/precomputed_panelAPP/GLOWgenes_prioritization_Retinal_disorders_GA.txt. Both scores were independently normalized using min-max normalization and equally weighted to generate a final normalized GDAS-GLOW score:

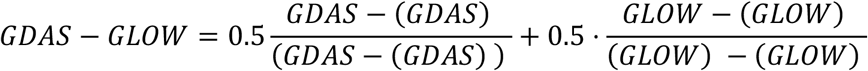

Molecular geneticists at the UH-FJD manually reviewed all variants identified in genes with prior evidence and data-supported candidate genes. For variants in genes without prior evidence or other genome regions, the review was restricted to the top 3,000 variants in homozygous state or in compound heterozygosity with another variant, sorted by their GDAS-GLOW score.

## RESULTS

### A Multi-Disease Local Database of Genomic Variant Frequencies

We built a local database of genetic variant frequencies (FJD-DB) from a cohort of 11,084 patients presenting a broad spectrum of genetic diseases, mostly rare disorders, from the Genetics and Genomics Department of the UH-FJD. For patients with multiple genetic tests, the NGS assay with the highest target coverage was selected for database integration. This cohort comprised Clinical Exome Sequencing (CES; ∼5,000 targeted genes; N=10,604; 95.7%), Whole Exome Sequencing (WES; N=354; 3.2%), and Whole Genome Sequencing (WGS; N=126; 1.1%) **(Figure 1A)**. At the sample level, the FJD-DB captured a median of ∼25,000 variants per CES, ∼270,000 variants per WES, and ∼4,700,000 variants per WGS. In total, the database aggregated 45,050,874 unique genetic variants, including single nucleotide variants (SNVs) and indels **(Figure 1B)**. Among these, 184,147 unique variants were classified as potentially pathogenic (see Methods).

**Figure 1.**
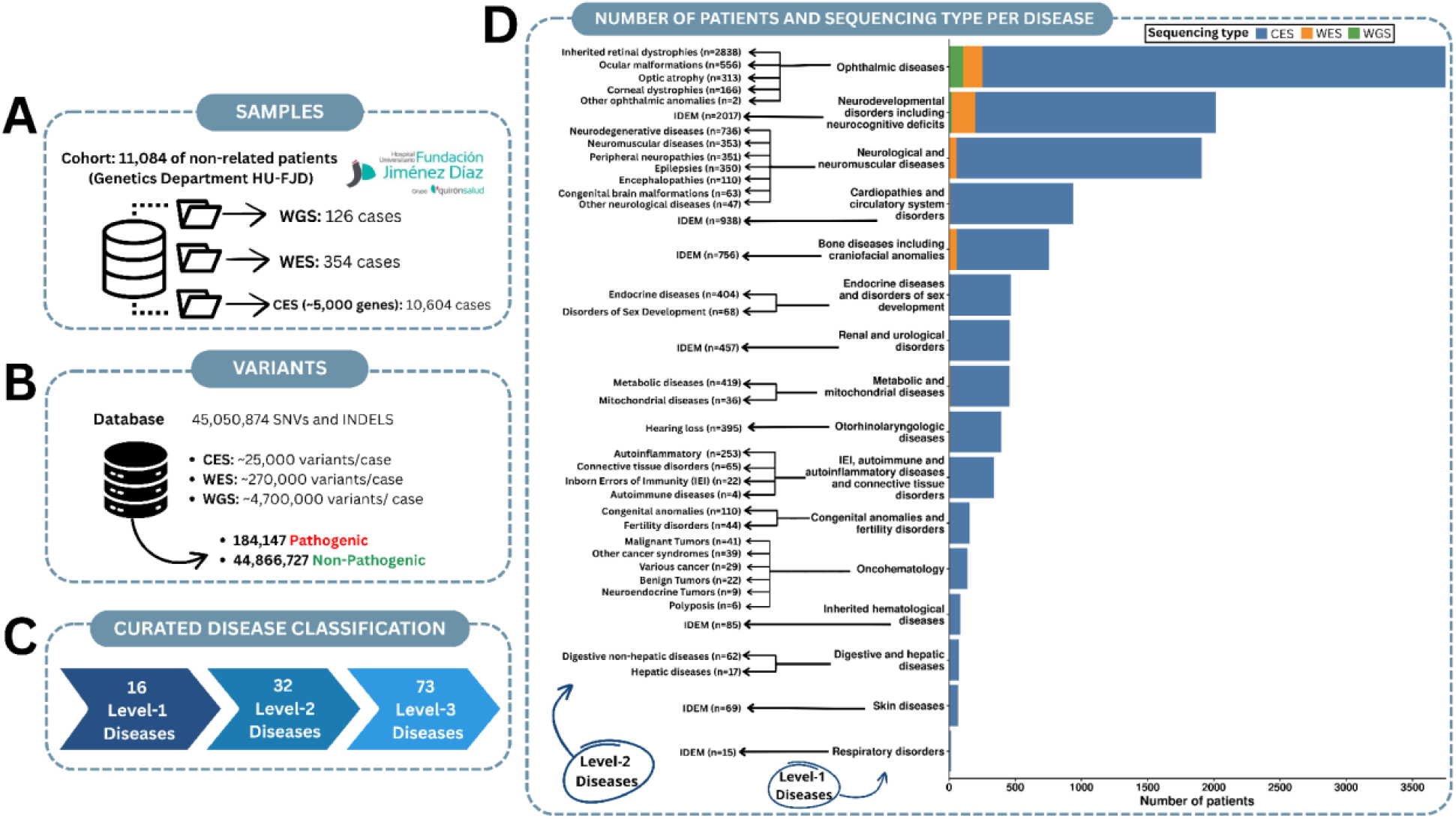
Overview of the FJD-DB. A multi-disease database with genetic variant frequencies from a cohort of 11,084 unrelated patients with suspected rare diseases. A) Distribution of patient samples and next-generation sequencing (NGS) assay types available in the database. B) Total number of unique genetic variants captured in the database, stratified by source, and clinical classification. C) Clinical classification of patients into three-hierarchical disease levels, from broad clinical categories (Level 1) to specific disorders (Level 3) after filtering. D) Quantitative breakdown of the patient cohort and sequencing approaches for the top-level disease categories (Level 1), further subdivided into their respective second classification (Level 2) clinical subgroups. Clinical Exome Sequencing, CES; Whole Exome Sequencing, WES; Whole Genome Sequencing, WGS).

Patients were clinically classified according to their primary diagnostic suspicion using a three-level hierarchical system (Levels 1 to 3), ranging from broader to more specific disease categories (Figure 1C, Supplementary Table S1). Each patient was annotated across the three hierarchical levels and could be assigned to multiple disease categories simultaneously. Among the Level-1 disease groups, Ophthalmic diseases comprised the largest number of patients (N = 3,750; 33.8%), followed by Neurodevelopmental disorders including neurocognitive deficits (NDD; N = 2,017; 18.2%), Neurological and neuromuscular diseases (N = 1,908; 17.2%), Cardiopathies and circulatory system disorders (N = 938; 8.5%) and Bone diseases including craniofacial anomalies (N = 756; 6.8%). Ophthalmic diseases and NDD also contain the highest number of patients sequenced by WGS or WES (226 and 199 samples, respectively) (Figure 1D).

### Characterization of Disease-Associated Pathogenic Variants

The FJD-DB was used to calculate genetic variant frequencies across patient subcohorts defined according to each clinical classification level. In parallel, variant frequencies were also determined for corresponding pseudocontrol subcohorts designed for each classification group. Next, we assessed the statistical association of each pathogenic variant present in the subcohorts defined by the Level-2 clinical classification using our Variant Disease Association Score (VDAS), which compares allele frequencies and homozygous counts between patients presenting with a given disease and their corresponding pseudocontrol subcohort (see Methods).

Figure 2 summarizes the main characteristics of variants showing a positive disease association (VDAS > 0) within each Level-2 disease subcohort compared with pseudocontrols, hereafter referred to as Disease-Associated Pathogenic Variants (DAPVs) (Supplementary Table S3). Some features correlate with the disease subcohort sample size. For instance, the total number of identified DAPVs increases with the number of patients (r = 0.98, p < 2.2 × 10^−16^), as well as the number of genes harboring at least one DAPV (r = 0.96, p < 2.2 × 10^−16^).

**Figure 2.**
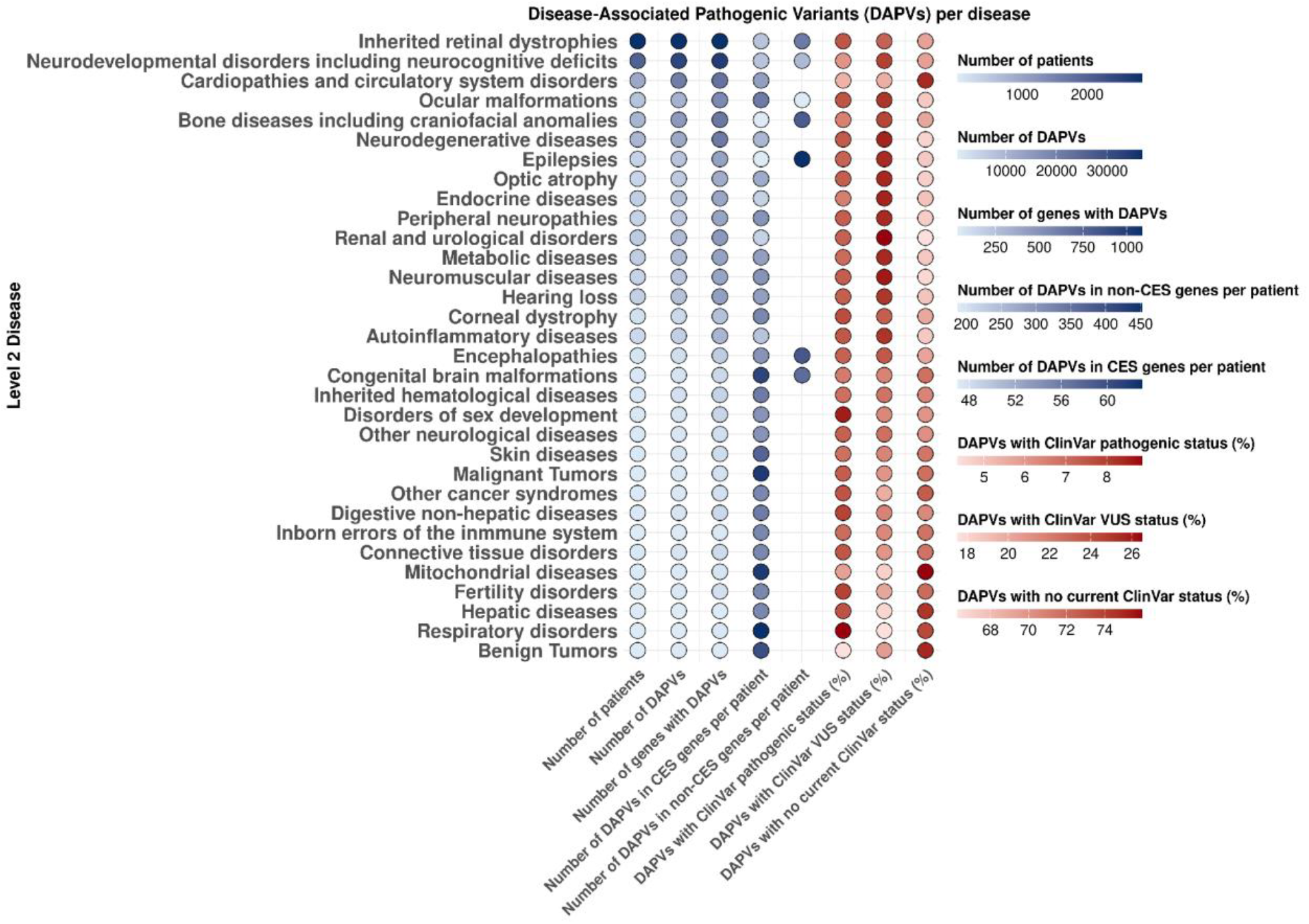
Characterization of Disease-Associated Pathogenic Variants (DAPVs) across disease subcohorts. Level-2 diseases are represented in every row.

Other features appeared to be independent of cohort size. This is the case of the DAPV burden per patient, both within genes included in CES panels (known gene-disease associations) and outside these panels (non-CES). Thus, Epilepsies and Bone diseases (including craniofacial anomalies) accumulated a relatively higher number of DAPVs in non-CES genes, whereas diseases such as Ocular malformations showed a stronger enrichment of DAPVs within CES genes.

Stratification of DAPVs according to their clinical significant annotations in the ClinVar database reveals highly heterogeneous behaviors across groups of diseases. The highest proportions of pathogenic or likely pathogenic DAPVs were observed in Respiratory disorders, Disorders of sex development and Fertility disorders, whereas Benign tumors and Cardiopathies and circulatory system disorders showed the lowest proportion. The latter two categories, together with Mitochondrial diseases, also had the highest proportions of variants lacking ClinVar annotations, while Neuromuscular diseases exhibited the highest proportion of VUS.

### Protein Domains Enriched for Disease-Associated Pathogenic Variants

For every Level-2 disease, predicted pathogenic variants were ranked according to their disease association strength as measured by their normalized VDAS (nVDAS). Gene Set Enrichment Analysis (GSEA) was subsequently used to identify protein domains enriched for these disease-associated variants. As variants were mapped to individual domains, these enrichments are domain-specific rather than gene-specific. In total, we identified 179 significantly enriched InterPro protein domains across 23 of the 32 diseases analyzed (see Methods for significance requirements). Most domains showed disease-specific enrichment, with 168/179 (93.9%) being significantly enriched in a single disease, whereas only 11/179 (6.1%) were shared between two diseases (Supplementary Table S4).

The number of enriched protein domains varies across diseases. While 21 of the 23 diseases showed between 1 and 14 enriched domains, NDDs and IRDs displayed a markedly higher number, with 71 and 52 enriched domains, respectively (Figure 3A), likely reflecting their larger proportion of patients sequenced by WES or WGS. The Top three protein domains enriched across Level-2 diseases are shown in Figure 3B.

**Figure 3.**
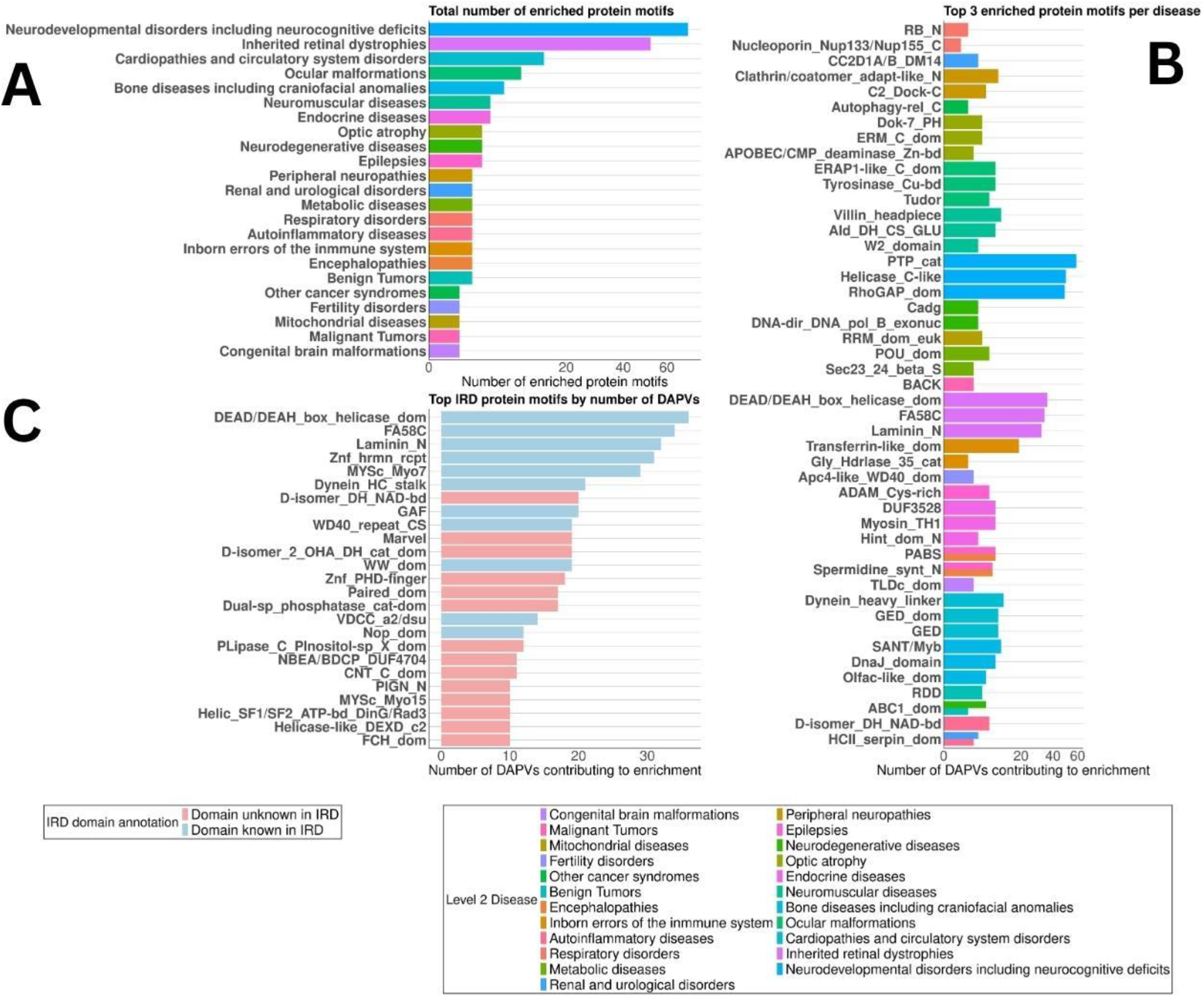
InterPro protein domains enriched for disease-associated pathogenic variants (DAPVs). A) Number of significantly enriched protein domains (InterPro short description naming) identified across Level-2 disease groups using Gene Set Enrichment Analysis (GSEA). Statistical significance was defined by False Discovery Rate (FDR) < 0.25; GSEA Normalized Enrichment Score (NES) > 0; >75% of the core enrichment variants within the enriched term had a normalized Variant Disease Association Score (nVDAS) > 0. B) Top three disease-specific enriched protein domains across diseases. Top domains were defined as those containing the highest number of leading-edge DAPVs contributing to the enrichment signal. C) IRD-enriched protein domains with ≥10 DAPVs, colored according to prior evidence linking the domain to the disease. Prior evidence is defined as the presence of at least one DAPV in an IRD-gene with prior evidence.

Focusing on IRDs, we distinguished between enriched domains containing DAPVs in at least one gene already associated with IRD, and domains exclusively in genes not previously linked to the disease. While the latter may highlight entirely novel disease-gene associations, the former may also point to additional candidate genes sharing disease-relevant functional domains (Figure 3C). Domains containing DAPVs mapped to genes already known to cause IRD included: MYSc_Myo7 (“Class VII myosin, motor domain”), DEAD/DEAH_box_helicase_dom (“DEAD/DEAH box helicase domain”), Dynein_HC_stalk (“Dynein heavy chain, coiled coil stalk”), FA58C (“Coagulation factor 5/8 C-terminal domain”), Laminin_N (“Laminin N-terminal”) or GAF (“GAF domain”). Additional domains enriched in IRDs were affected by DAPVs in genes that have not yet linked to IRDs, including MYSc_Myo15 (“Class XV myosin, motor domain”) and Helicase-like_DEXD_c2 (“Helicase-like, DEXD box c2 type”), potentially suggesting novel candidate genes for IRDs (Figure 3C).

### Genes Enriched in Disease-Associated Pathogenic Variants

To identify genes with a high mutational burden of variants associated with specific diseases, we developed the Gene Disease Association Score (GDAS). The GDAS represents the relative accumulation of pathogenic variants within a gene in a specific disease cohort compared to its corresponding pseudocontrol cohort. For each Level-2 subcohort, we define High Pathogenic Burden Genes (HPBGs) as those showing an increased pathogenic mutational burden in cases than in pseudocontrols (GDAS > 0; Supplementary Table S5). Most HPBGs were disease-specific, with 222/242 (91.7%) identified exclusively in a single disease, whereas only 20 genes (8.3%) were shared between two distinct disorders.

To explore the strongest HPGB-disease associations (GDAS>3 and highest aggregated DAPV counts), we classified genes into two categories: i) known (“in-panel”) genes, as those previously associated with the disease (Supplementary Table S2), and ii) potentially novel (“out-of-panel”) genes, as those not currently linked to pathogenesis of that specific disease. Among the top known genes across diseases (Figure 4, blue tiles), those associated with IRDs showed the highest aggregated DAPV counts (*ABCA4, USH2A, CRB1, BEST1*, and *PRPH2*). Other notable examples include *CATSPER* and *GJB2* associated with Hearing loss, *TGFB1* with corneal dystrophy, *OPA1* with optic atrophy, and *COL4A4* with Renal and urological disorders. Among the top novel genes across diseases, the highest aggregated DAPV counts were observed for *MUC3A* and *SERPINA1*, both identified among the top five out-of-panel genes in IRDs and Ocular malformations. Similarly, *HLA-DRB1* appeared among the top five novel genes in NDDs, while also being classified as a top in panel gene in Peripheral neuropathies. Although showing lower aggregated DAPV counts, *GLP1R* was also identified among the top-five novel genes in two diseases: Corneal dystrophy and Metabolic diseases.

**Figure 4.**
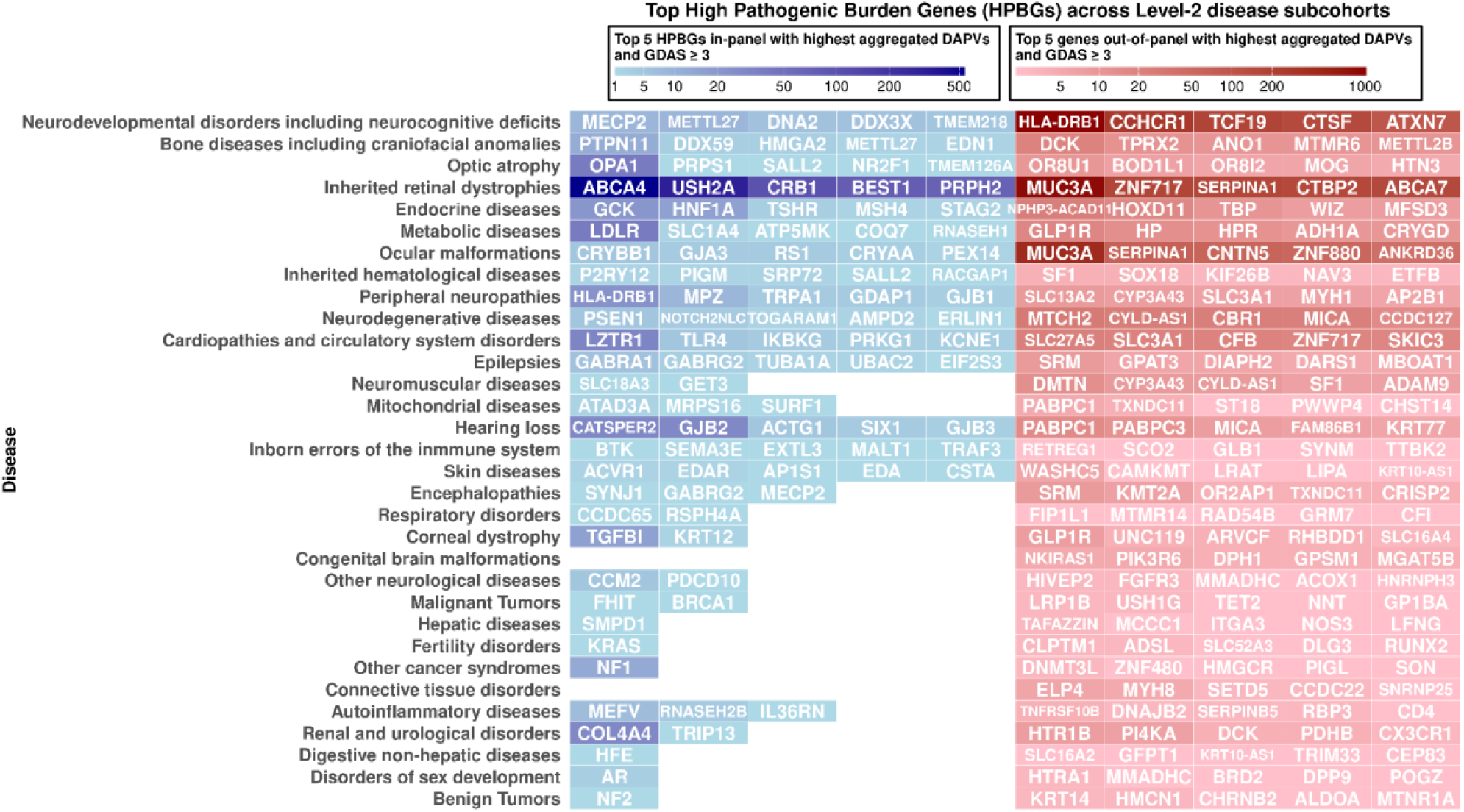
Top High Pathogenic Burden Genes (HPBGs) across Level-2 disease subcohorts. The matrix illustrates the top five known genes (“in-panel”, represented in blue tiles) and the top five potentially novel genes (“out-of-panel”, represented in red tiles) prioritized for each Level-2 clinical subcohort according to the Gene Disease-Association Score (GDAS > 3) and aggregated Disease-Associated Pathogenic Variants (DAPV) counts in patients. The color intensity gradient represents the magnitude of the aggregated DAPVs and the corresponding GDAS, with darker intensity indicating more robust disease-association signals.

### Disease-Specific Functional Landscape of High Pathogenic Burden Genes

To explore the biological functions most affected by pathogenic variants across Level-2 diseases, we performed a GSEA using all genes ranked according to their normalized GDAS values (see Methods). This analysis enabled the identification of enriched HPO terms, biological pathways (KEGG and Reactome), and biological processes (Gene Ontology, GOs) in every Level-2 disease (Supplementary Table S6).

From the HPO findings, we identified 124 significantly enriched terms distributed across 13 of the 32 analyzed diseases (Figure 5A). Remarkably, almost all enriched HPO terms were disease-specific, with only three terms shared between two distinct diseases. The clinical subcohorts with the highest number of enriched HPO terms were IRDs (N = 41) and Cardiopathies and circulatory system disorders (N = 34). In IRDs, most enriched terms were directly related to retinal phenotypes (“Pigmentary retinopathy”, “Undetectable electroretinogram”, and “Retinal atrophy”). Other enriched terms were related to abnormalities of metabolism/homeostasis and the endocrine system (“Hyperinsulinemia”, “Abnormal circulating insulin concentration”, and “Maturity-onset diabetes of the young”). In Cardiopathies and circulatory system disorders, enriched HPO terms spanned 10 different phenotype categories, prominently including “Abnormality of the cardiovascular system”.

**Figure 5.**
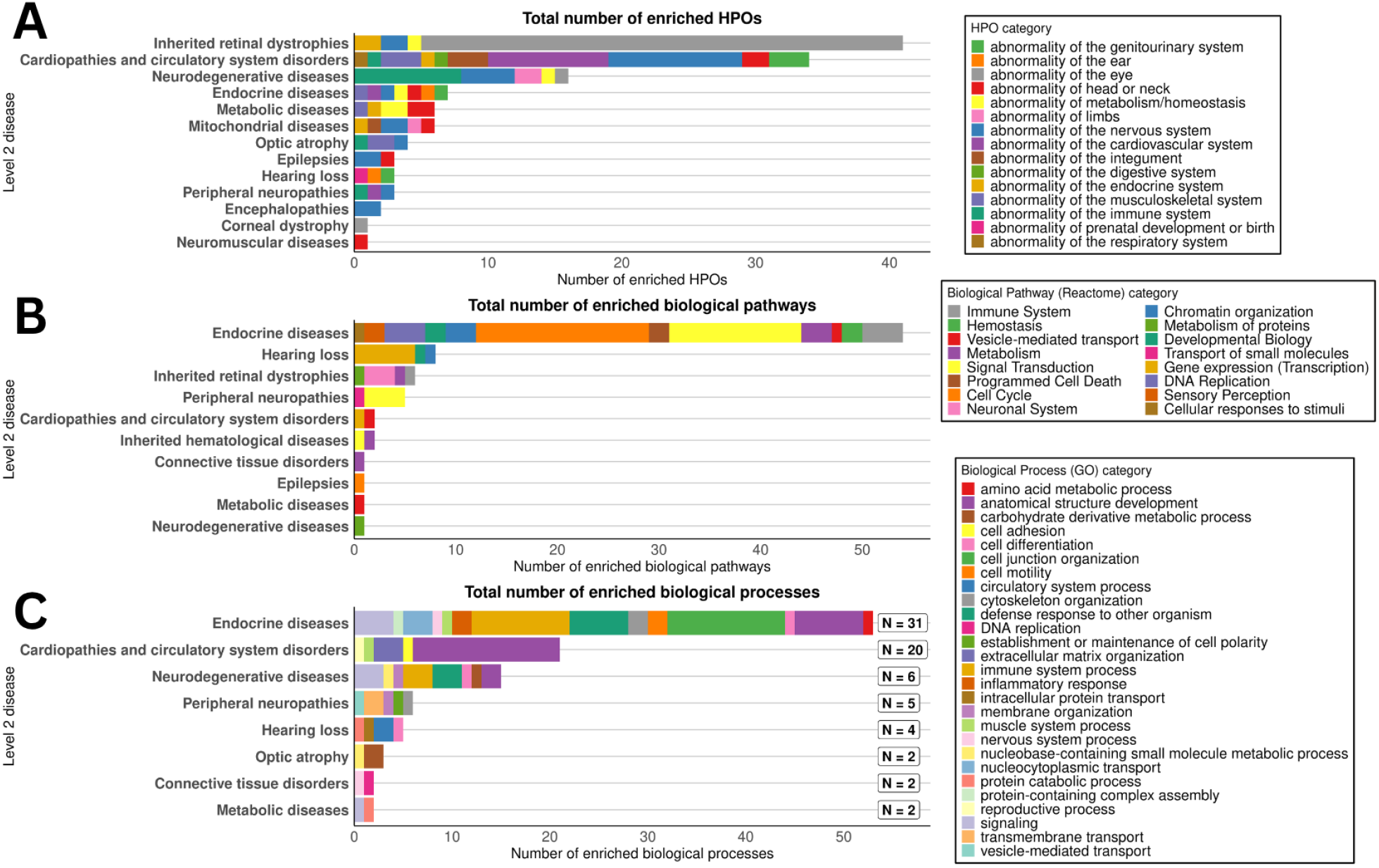
Disease-specific functional profiles of High Pathogenic Burden Genes (HPBGs) across Level-2 disease groups. Gene Set Enrichment Analysis (GSEA) profiles for HPBGs, classified by database source: A) Human Phenotype Ontology (HPO) terms, B) Reactome pathways, and C) Gene Ontology (GO) biological processes. Only significantly enriched terms assigned to their higher-level functional category are shown. Statistical significance was established by False Discovery Rate (FDR) < 0.25; GSEA Normalized Enrichment Score (NES) > 0; and >75% of the core enrichment genes within the enriched term had a normalized GDAS (nGDAS) > 0). As individual GO terms can belong to multiple higher-level categories, these counts are not unique, and a single term may contribute to multiple categories. The square marker denotes the number of uniquely enriched GO terms associated with each Level-2 disease subcohort.

Regarding the biological pathways from the Reactome database, we identified 79 significantly associated terms in 10/32 diseases. Of all, 78 (98.7%) were disease-specific and only a single pathway (1.3%), “Glucuronidation”, was enriched in three diseases: Connective tissue disorders, Endocrine diseases and Inherited hematological diseases (Figure 5B). We also identified eight significantly enriched terms from the KEGG database in 5/32 diseases (Supplementary Table S6).

In terms of the biological processes, we identified 72 significant GO terms in 8/32 diseases. Most of the terms are uniquely linked to a single disease except for four GO terms (5.55%), all related to metabolism. Thus, three metabolic terms (“Cellular glucuronidation”, “Glucuronate metabolic process” and “Uronic acid metabolic process”) were enriched in Connective tissue disorders, Hematological and Endocrine diseases, and term “Flavonoid metabolic process”, was enriched in those three diseases and also in Peripheral neuropathies (Figure 5C).

### Data-Guided Reanalysis of Unsolved Inherited Retinal Dystrophies Cases

Building on these results, we designed a diagnostic algorithm (Figure 6) to support a guided reanalysis of unsolved patients within a cohort of a specific RD. This framework integrates VDAS, GDAS and functional enrichment knowledge information to filter and prioritize variants. As proof of concept, this algorithm was applied to the unsolved IRD cases within our FJD-DB. Starting from over 1,100 unsolved IRD cases, we extracted a total of 112,378 variants pathogenic or predicted pathogenic, rare in the general population (gnomAD AF<0.01) and showed a higher frequency in IRD cases compared to pseudocontrols (VDAS>0). These variants were classified into three groups according to the level of prior clinical evidence linking their corresponding genes to IRDs: 1) variants in genes with prior evidence, that included 11,520 variants, 2) variants in data-supported IRD candidate genes, comprising 1,150 variants, and 3) variants located in genes or genomic regions not previous linked to IRDs, that encompassed 99,708 variants. To prioritize this latter group, variants were ranked using a combined metric (GDAS-GLOW) based on both GDAS values with the gene’s corresponding score from GLOWgenes, a gene-disease predictor algorithm optimized for rare diseases [35]. We further focused on variants compatible with a recessive mode of inheritance, specifically those found in homozygous or potential compound heterozygosity states (23,990 variants).

**Figure 6.**
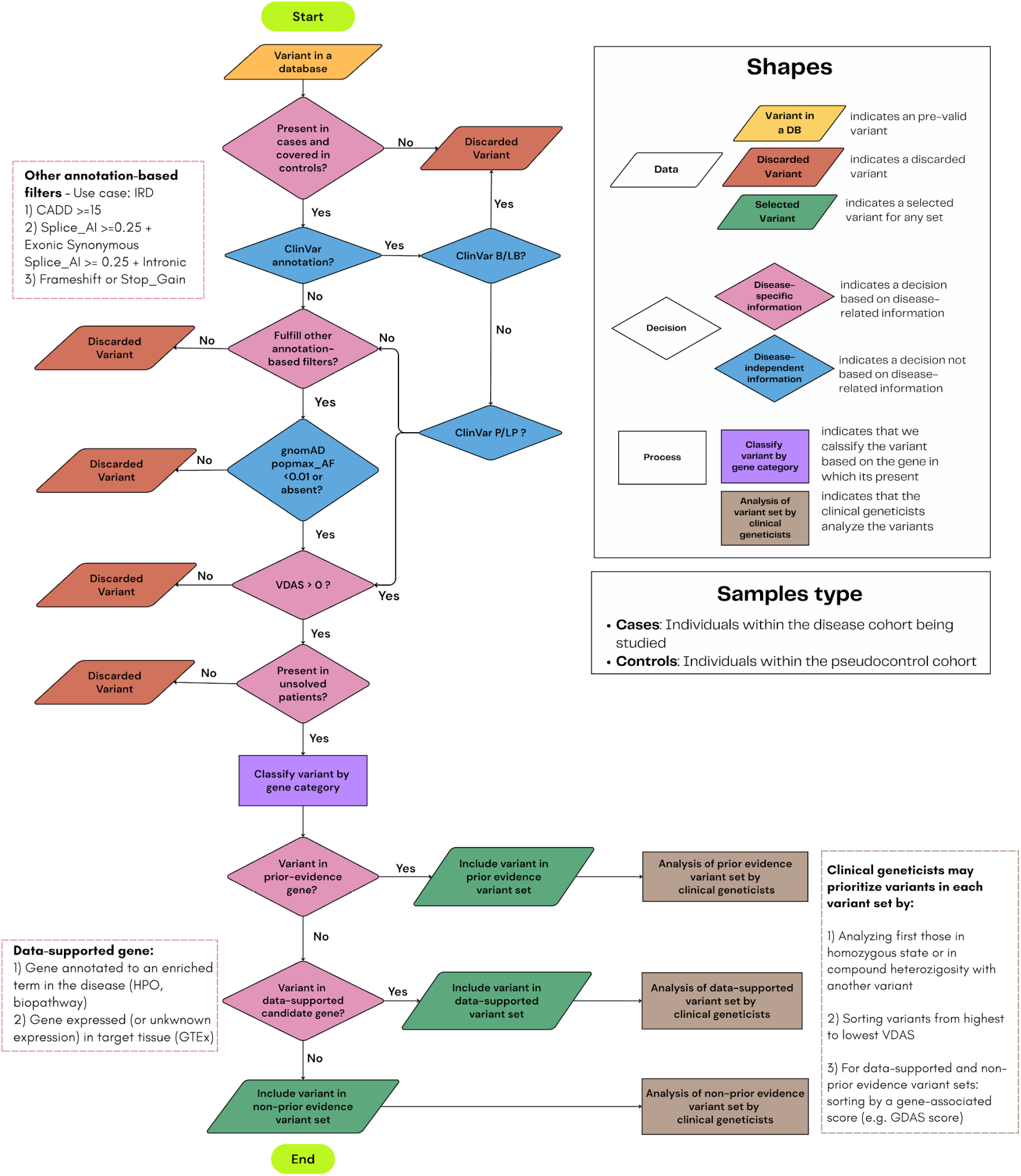
A data-driven diagnostic framework for the guided reanalysis of unsolved rare disease cases using aggregated clinical genomic data.

All prioritized variants in genes with prior evidence and in data-supported IRD candidate genes were manually reviewed by clinical geneticists from the HU-FJD Genetics Department in the context of the corresponding patients’ clinical records. From the variants in homozygous and in compound heterozygosity found within genes without prior evidence or other genome regions, the first 3,000 prioritized variants, sorted by their GDAS-GLOW score, were also selected for clinical review.

Overall, we report 36 variants that provided diagnostic evidence for 32 previously unsolved IRD cases (Table 1). Among these, eight variants were known associated variants located in IRD prior-evidence genes but had been overlooked at the time of the patients’ initial analysis. We additionally identified 20 novel variants, of which 18 were classified as VUS and two as likely pathogenic in IRD-prior evidence genes. We further highlighted four novel variants in two data-supported candidate genes (*DMXL2* and *KIF21A*) and three novel variants in two potential candidate genes prioritized using the combined GDAS-GLOWgenes score (*DNM1* and *NAGLU*). In total, three patients received a definitive molecular diagnosis and 25 achieved a molecular diagnosis pending additional testing (possibly solved). Additionally, we propose four new candidate genes which provide potential new insights into the molecular basis of IRD.

**Table 1.**
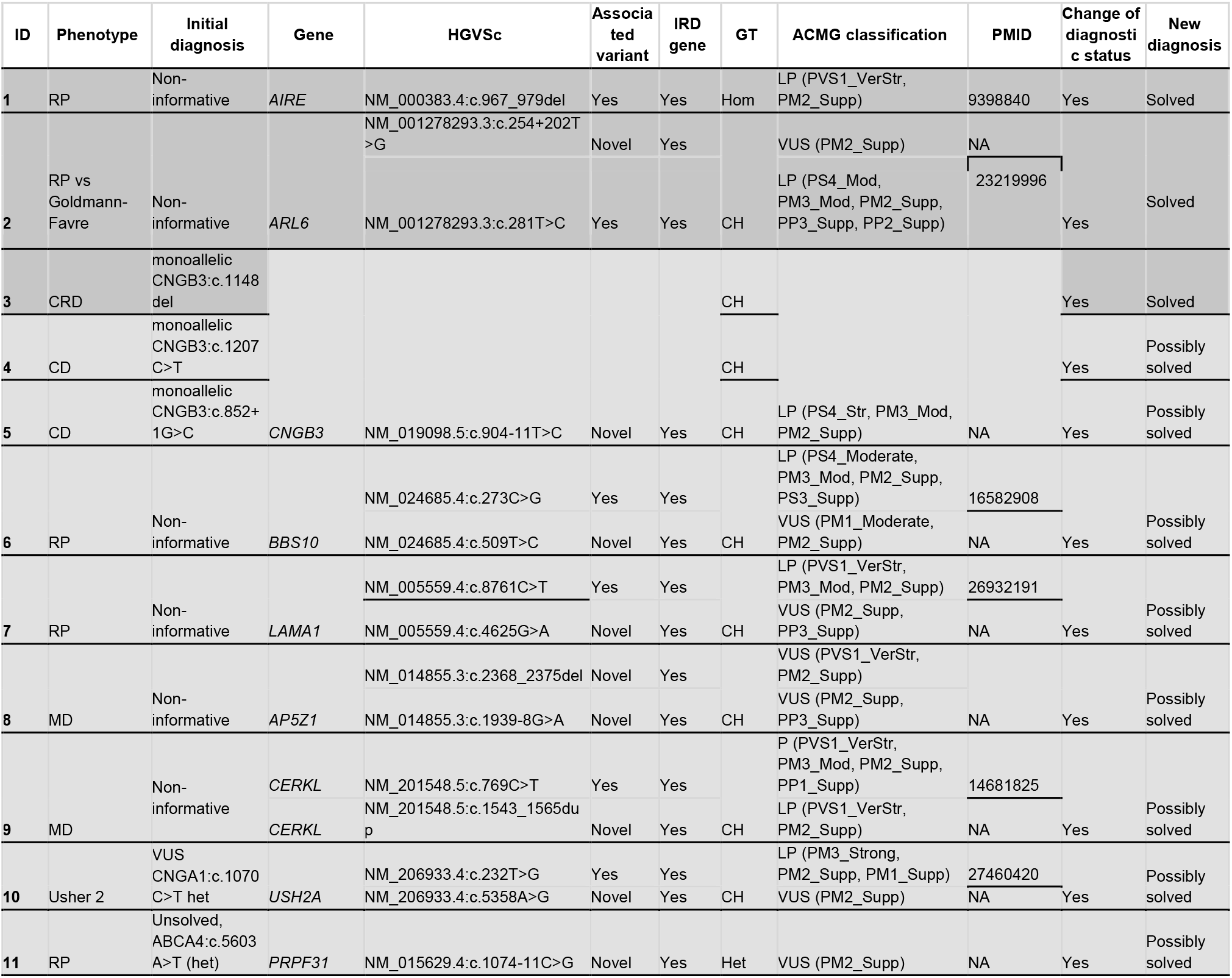
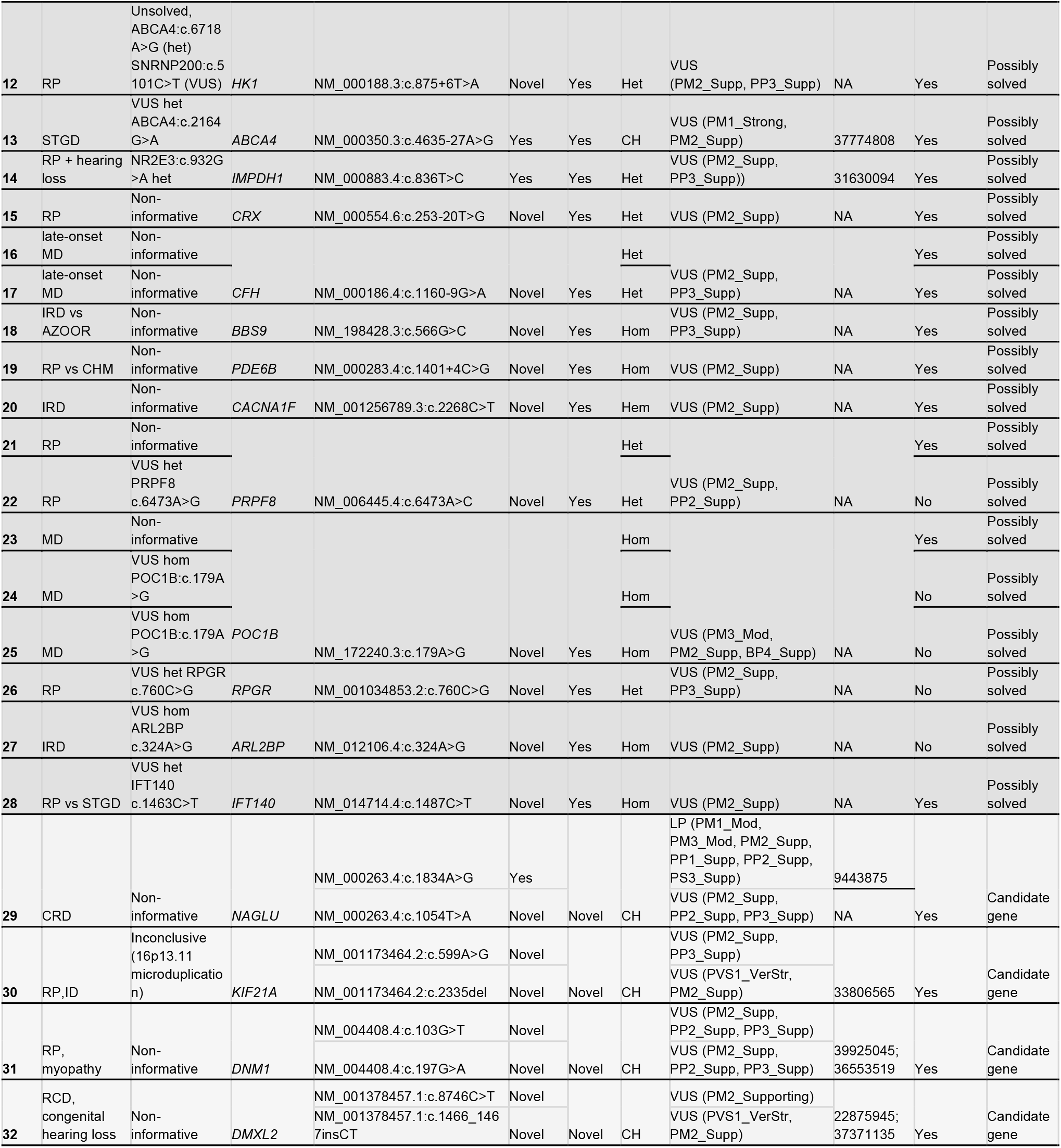
Genetic variants providing additional molecular evidence in previously unsolved Inherited Retinal Dystrophies (IRD) cases. For each variant, the corresponding patient is described, including its: ID, phenotype (RP= retinitis pigmentosa, CRD=cone-rod dystrophy, CD= cone dystrophy, STGD= Stargardt disease, MD=macular dystrophy, ID=intellectual disability) initial and revised diagnoses, change of diagnostic status (no=initial and revised diagnoses are the same, yes=initial and revised diagnoses are different) and genotype (Het = Heterozygous, Hom = Homozygous, CH = Compound heterozygous, Hem = Hemizygous). Variant details include Human Genome Variation Society coding sequence nomenclature (HGVSc), novelty, American College of Medical Genetics and Genomics (ACMG) classification (initial and new), the affected gene and its IRD-association status (yes = IRD prior-evidence gene, novel = new-gene phenotype association) and relevant PMID references. ACMG strength evidence is abbreviated as follows: Supp = Supporting, Mod = Moderate, Str = Strong, VerStr = Very Strong. ACMG Classification is abbreviated as follows: LP= Likely pathogenic; VUS= Variant of Unknown Significance).

## DISCUSSION

There is a growing global call to reduce the diagnostic odyssey experienced by patients with RDs [3]. In this work, we demonstrate the utility of real-world sequencing data as a powerful engine for disease mechanism discovery. We generated disease-specific variant and gene prioritization frameworks that contributed to leverage the mutational burden across more than 11,000 patients to identify disease-associated protein domains, novel candidate genes, and molecular functions and pathways specifically altered across 32 disease groups.

First, we introduce VDAS, a novel computational framework for prioritizing genetic variants through case-control comparisons. While *in silico* pathogenicity predictions are inherently uncertain, aggregating signals across genes, protein domains and biological functions reduces stochastic noise to uncover consistent disease-associated patterns. As expected, the number of prioritized variants was strongly influenced by subcohort sample size and sequencing target coverage. However, comparative analysis across clinical categories also revealed differences in the underlying genetic architecture and current disease knowledge that were not explained by either factor. For instance, the Epilepsies and Encephalopathy cohorts showed a significant enrichment of prioritized variants in genes not yet linked to human disease, suggesting substantial undiscovered genetic causes. In contrast, Mitochondrial diseases and Cardiopathies and circulatory system disorders showed lower proportions of clinically classified variants, whereas Peripheral neuropathies, Epilepsies, Neurodegenerative, and Neuromuscular diseases accumulated the highest percentages of VUS. Together, these patterns provide a data-driven view of remaining knowledge gaps.

We further used VDAS to identify the accumulation of potentially pathogenic variants in protein domains, genes, biological processes, and molecular pathways in disease cases relative to pseudocontrols.

Regarding protein domains, subcohorts with the highest genomic coverage (IRD, NDD) showed the largest number of associated protein domains, driven by the higher number of genes tested in exome- and genome-wide approaches. We used IRDs as a case study to illustrate the findings obtained from the protein domain analysis. For instance, the GAF domain (IPR003018) [36,37] accumulated DAPVs in *PDE6A, PDE6B*, and *PDE6C*, all well-established IRD genes [38–41], as well as in *PDE11A*, which has not yet been associated with IRDs but expressed in retina according to GTEx and with a mutation reported linked in a reduced risk of developing myopia in humans case [42]. Similarly, the Class VII myosin motor domain (IPR036106) was significantly enriched in variants affecting both *MYO7A*, the canonical cause of Usher syndrome type 1B [43], and *MYO7B*, a related gene recently proposed as a candidate for retinitis pigmentosa [44]. The Dynein heavy chain coiled, coil stalk (IPR024743) also has a significant pathogenic mutational burden in the known IRD ciliary gene *DYNC2H1* [45], as well as in unassociated genes such as *DYNC1H1*, which shares highly structural homology with *DYNC2H1 [46]*, and has been implicated in retinal degeneration in experimental animal models [47–49], or *DNAH14* which has been reported among the predicted targets of dysregulated miRNAs in degenerating canine retinas [50], suggesting a potential involvement in retinal disease-associated regulatory networks. Likewise, the DEAD/DEAH box helicase domain (IPR011545) was enriched in DAPVs within the IRD-associates *SNRNP200* gene [51] and *DDX41*, recently associated to IRDs [52] and other genes not yet linked to IRDs, such as: *RECQL4*, whose pathogenic variants cause Rothmund-Thomson syndrome, a disorder with reported retinal manifestations [53] or *DDX25*, candidate gene for color agnosia [54]. A final example among domains enriched in established IRD genes is the Laminin N-terminal domain (IPR008211), which accumulated DAPVs in several known IRD genes (*LAMA1* [55], *LAMB2 [56]* or *USH2A [57]*) and in others not yet associated, such as *LAMA2*, associated with retinal ganglion cell outgrowth during development and further detected in the adult mouse [58], *LAMA5*, with a mutation associated to early posterior vitreous detachment described [59] or *LAMC3* which is implicated in retina angiogenesis and astrocyte migration [60]. Other significant enriched domains were identified exclusively in genes not currently associated with IRDs. This is the case of the Helicase-like, DEDX box c2 type domain (IPR006554), which is present in four proteins genome-wide. We observed a significant accumulation of DAPVs affecting this domain in two of these proteins, ERCC2 and RTEL1. Neither of which are encoded by an IRD gene, but both show biological connections to retinal maintenance. ERCC2, a subunit of the transcription factor IIH (TFIIH), associated with genomic instability syndrome binds to the promoter and coding regions of opsin genes together with the photoreceptor transcription factors CRX/NR2E3 [61], both well-established IRD genes [62,63]. Similarly, vitreoretinopathy has been reported in two patients carrying rare variants in *RTEL1 [64,65]*, related to a short telomere syndrome. These findings demonstrate that domain-level burden analysis can uncover hidden potential pathological associations that are missed by single-gene filters. Additional insights may be obtained from the remaining enriched domains, which are provided as a resource for the community (Supplementary Table S4).

We next explored the genes with the highest pathogenic mutational burden in cases compared to pseudocontrols in each disease group. Here, we observe known gene-disease associations with strong statistical support. For instance, in the IRDs subcohort, *ABCA4, USH2A, CRB1, PRPH2* and *BEST1* ranked in the top five known genes, directly reflecting the known most prevalent causes within our cohort [28], one of the largest worldwide. This validation was replicated across other clinical subgroups: among the top five genes, *GJB2* emerged in Hearing loss [66], *COL4A4* in Renal and urological disorders [67], *OPA1* in optic atrophy [68], and *TGFBI* in corneal dystrophy [69].

The primary translational value of this data-driven framework lies in its capacity to prioritize highly ranked, out-of-panel genes as potential novel disease candidates to be further explored. For instance, *MUC3A* emerged as a top non-associated gene in both IRD and Ocular malformations subcohorts. While *MUC3A* itself is not currently known to be expressed in ocular tissues [70], multiple members of the mucin family play essential roles in maintaining tear film integrity and ocular surface homeostasis [71–73]showing that the enrichment observed in *MUC3A* may reflect previously unexplored mechanisms related to ocular disease. Likewise, *SERPINA1*, also enriched in IRDs and Ocular malformations, exhibits altered levels across multiple ophthalmic conditions. It is known to be upregulated in the aqueous humor of glaucoma, cataract, and high myopia patients, and in the vitreous fluid of patients with age-related macular degeneration but reduced in the corneal epithelium and stroma in keratoconus and in the serum of diabetic retinopathy patients [74].

In the NDD subcohort, the out-of-panel gene with the highest pathogenic mutational burden was *HLA-DRB1*. This major histocompatibility complex locus is increasingly being studied for its relationship with Autism Spectrum Disorders [75,76], language impairment or attention deficit hyperactivity disorder [77]. We further highlighted *GLP1R* as a top novel candidate across two highly distinct categories: Metabolic diseases and Corneal dystrophy. While polymorphisms in *GLP1R* are associated metabolic traits, such as accelerated body mass index growth during development, insulin resistance and higher compensatory insulin secretion [78], recent clinical evidence indicated that therapy based on GLP1R receptor agonist (GLP1R-RA) shows a potential protective, anti-inflammatory effects on the ocular surface of patients with diabetes [79]. These examples suggest that gene’s DAPV enrichment analyses may identify not only novel monogenic candidate genes, but also may uncover genetic modifiers and biological factors contributing to clinical penetrance, disease susceptibility, progression, or phenotypic variability.

Biological processes and molecular pathways represent the highest level of biological organization explored in our analyses. By identifying biological processes and pathways enriched in DAPVs, we sought to uncover the functional landscape altered in each disease group and gain insights into the molecular mechanisms shaping their genetic architecture. Thus, these findings provide a source of novel biological hypotheses for future investigation. For instance, in IRDs, several enriched pathways were related to mucin biology and O-glycosylation (“Defective *GALNT3* causes HFTC”, “Defective *C1GALT1C1* causes TNPS”; “Dectin-2 family”, “Defective *GALNT12* causes CRCS1” and “Termination of o-glycan biosynthesis”), primarily driven by variants in multiple mucin genes. As mentioned before, mucins and eye related processes have already been linked and additional evidence suggests that cone opsins undergo mucin-type O-glycosylation mediated by GALNT enzymes, and disruption of these modifications leads to photoreceptor degeneration and altered retinal function in animal models [80]. Together, these observations suggest that alterations in mucin-related glycosylation pathways may represent a previously underappreciated mechanism contributing to retinal disease.

Finally, we wrapped all resources generated in this work into a data-driven reanalysis framework designed to translate cohort-level discoveries into patient-level diagnoses. Unlike conventional reanalysis approaches, which are guided by existing disease knowledge and predefined gene panels, our framework prioritizes a small list of candidate variants per patient based on evidence derived from the aggregated analysis of the cohort. As a result, hidden variants can be identified not only because they occur in known disease genes, but also because they are supported by disease-specific signals in currently-not-associated-to the disease genes. Overall, and only applied to IRDs due to resource constraints, dozens of patients were able to benefit from this exercise. As all our prioritizations are available, external laboratories can readily apply this clinical algorithm to the diagnosis of their own unsolved RD cases. In addition, we provide software to build the database and implement the proposed analytical framework.We acknowledge some limitations intrinsic to both the use of real-world clinical data and rare disease research. First, several disease subcohots are inherently small, restricting the statistical power required to detect robust disease-specific associations. The major limitation, however, resides in the uneven target NGS coverage across different disease subgroups. While some subcohorts groups benefit from an extensive WES and WGS coverage, most historical NGS data resides within targeted commercial CES panels, limiting the potential for discovering variants in non-coding or in out-of-panel genes. Nevertheless, this study was conceived as an exploratory effort designed to maximize the use of real-world diagnostic data across a broad spectrum of rare disease, including subcohorts with modest sample size. Taken together, our findings indicate that the aggregated nature of the analyses, together with the use of pseudocontrol-based comparisons, increases the likelihood that the identified enrichments reflect genuine underlying biological signals, whether directly related to disease causality or to broader disease-associated phenotypes and modifier effects [81]. Clinical patient categorization was guided by the expertise of clinical geneticists at a single center with the aim of maximizing the framework’s immediate utility for diagnosing future patients. Still, adopting automated HPO-based phenotyping or other standardized classification approaches may offer a more universally scalable strategy. Finally, due to the enormous volume of data generated across the 32 medical specialties, we have limited our in-depth clinical interpretation to our main area of expertise (IRD). Consequently, some findings in other disease groups may require more specialized evaluation and disease-specific refinement of the analytical framework. In particular, variant filtering and prioritization criteria optimized for IRD may not perfectly capture the genetic architectures of other disorders, and future studies will benefit from tailoring these parameters to the specific disease group.

## CONCLUSION

This study demonstrates that routine clinical genomic data can be repurposed to generate novel biological hypotheses, prioritize candidate variants and genes, characterize disease-specific functional landscapes, and support the reanalysis of unsolved cases. As clinical genomic datasets continue to grow globally, the implementations of computational frameworks such as the one presented here may will improve diagnostic rates and accelerate variant and gene discovery across the spectrum of rare diseases.

## Data Availability

All data generated or analyzed during this study are included in this published article and its Supplementary Information.

## DECLARATIONS

### Ethics approval and consent to participate

The project was reviewed and approved by the Research Ethics Committee of UH-FJD (Ref. 2023/06) and fulfills the principles of the Declaration of Helsinki and subsequent reviews. All samples included in this work were pseudonymized, and genomic data were only treated in aggregation. All patients individually studied signed an informed consent before participating.

## Consent for publication

Not applicable

## Authors’ contributions

PM and GUR conceived and designed the study. GUR, PM, YB designed computational analyses. GUR, PM, LFC, ALA analyzed the data, MJTT, AO, MC, AAF, BA, CA designed clinical classification and classified patients. LFC, ALA, CR, LLL, CA contributed to the interpretation of the results. GUR and PM wrote the paper, and all authors participated in the review and revision of the manuscript. PM conceived the original idea and supervised the project. All authors read and approved the final manuscript.

## Acknowledgements

We thank the patients for consenting to the use of their data for the study. We also thank all technical staff in the Genetics Department of the UH-FJD for conducting the sequencing and further analysis.

## List of abbreviations

RD: Rare diseases
FJD-BD: multi-disease genomic variant frequency database
UH-FJD: University Hospital - Fundacion Jiménez Díaz
IRD: Inherited Retinal Dystrophies
NGS: Next Generation Sequencing
CES: Clinical Exome Sequencing
WES: Whole Exome Sequencing
WGS: Whole Genome Sequencing
AF: Allele Frequency
AC: Allele Count
AN: Allele Number
VDAS: Variant Disease Association Score
GSEA: Gene Set Enrichment Analysis
nVDAS: Normalized Variant Disease Association Score
FDR: False Discovery Rate
NES: Normalized Enrichment Score
GDAS: Gene Disease Association Score
HPBGs: High Pathogenic Burgen Genes
nGDAS: Normalized Gene Disease Association Score
ERDC: European Retinal Disease Consortium
SNVs: Single Nucleotide Variants
NDD: Neurodevelopmental disorders including neurocognitive deficits
DAPVs: Disease-Associated Pathogenic Variants
VUS: Variant of Unknown Significance
HPO: Human Phenotype Ontology term
GO: Gene Ontology term

